# Time-Varying Effectiveness of Three Covid-19 Vaccines in Puerto Rico

**DOI:** 10.1101/2021.10.17.21265101

**Authors:** Mónica M. Robles Fontán, Elvis G. Nieves, Iris Cardona Gerena, Rafael A. Irizarry

## Abstract

**Background:** As of October 1, 2021 2,217,547 individuals were fully vaccinated against COVID-19 in Puerto Rico. Since the vaccination process commenced on December 15, 2020 111,052 laboratory-confirmed SARS-CoV-2 infections have been reported. These data permitted us to quantify the benefits of the immunization campaign and to compare effectiveness of the mRNA-1273 (Moderna), BNT162b2 (Pfizer), and Ad26.COV2.S (J&J) vaccines.

**Methods:** Department of Health databases holding vaccination status, SARS-CoV-2 test results, and COVID-19 hospitalizations and deaths were integrated. We fit a statistical model that adjusted for time-varying incidence rates and age to estimate vaccine effectiveness and hospitalization and death relative risks. Code and data are provided here: https://github.com/rafalab/vax-eff-pr.

**Results:** At the peak of their protection, mRNA-1273, BNT162b2, and Ad26.COV2.S had an effectiveness of 90% (88%-91%), 87% (85%-89%), and 58% (51%-65%), respectively. After four months, effectiveness waned to about 70%, 60%, and 30%. We found no evidence that effectiveness was different after the Delta variant became dominant. For those infected, the vaccines provided further protection against hospitalization and deaths across all age groups. All vaccines had a lower effectiveness for those over 85 years, with a larger decrease for the Ad26.COV2.S vaccine. Overall, thousands of hospitalizations and deaths were avoided thanks to the vaccines.

**Conclusions:** The mRNA-1273 and BNT162b2 vaccines were highly effective across all age groups. They were still effective after four months although the protection waned. The Ad26.COV2.S vaccine was effective but to a lesser degree, especially for older age groups.

---

The first COVID-19 case was first reported on December 31, 2019. The race for a vaccine commenced shortly after. By fall 2020, there were several ongoing clinical trials for vaccines developed with full messenger RNA (mRNA) technology.^1^ In the United States, three of the currently used COVID-19 vaccines were shown to be highly effective against infection, hospitalizations, and deaths in randomized double-blind trials. A two-dose series of BNT162b2 (Pfizer) COVID-19 vaccine was shown to be 95% efficacious against symptomatic COVID-19 in individuals 16 and older.^2^ Similarly, a two-dose series of mRNA-1273 COVID-19 provided 94% efficacy against symptomatic COVID-19 for those 18 and older.^3^ Finally, a single-shot of the Ad26.COV2.S COVID-19 vaccine provided 66% protection for individuals 18 years and older.^4^ These findings led to emergency use approvals (EUA) by the Federal Drug Administration (FDA) for these three vaccines. Over 380 million doses of these vaccines have been administrated in the United States.

Since the start of the vaccination process, the effectiveness of the COVID-19 vaccines has been estimated in several observational studies. A population-based study performed early during the vaccination process in Israel estimated effectiveness to be 72% for the BNT162b2 vaccine.^5^ A case-control study, also in Israel, found 87% effectiveness for the BNT162b2 vaccine, however, lower protection against hospitalizations and deaths.^6^ These estimates have been questioned due to lack of adjustment for age. A study conducted in inpatient care settings, across 187 hospitals in the United States, compared all three vaccines found the mRNA-1273, BNT162b2, and Ad26.COV2.S COVID-19 vaccines were found to be highly effective against hospitalization and death.^7^ Other observational studies have estimated vaccine effectiveness in the midst of variants of concern. The effectiveness against infection in the dominance of the Delta (B.1.617.2) variant has been shown to remain at 88% for the BNT162b2 COVID-19 vaccine.^8^ Similarly, in a population-based observational study found that two doses of the mRNA-1273 and BNT162b2 COVID-19 vaccines provided substantial protection against the Alpha (B.1.1.7), Beta (B.1.351), Gamma (P.1), and Delta variants of concern.^9^ Nonetheless, real-world population based estimates of the mRNA-1273, BNT162b2, and Ad26.COV2.S COVID-19 vaccines effectiveness against infections, hospitalizations and deaths is still limited. Here we add to these findings by examining the effect of vaccines on all laboratory-confirmed SARS-CoV-2 infections observed in Puerto Rico since the vaccination process started, which provided enough power to estimate effectiveness across age groups and as function of time since the individuals are considerd fully vaccinated (14 days after final dose).

Following the emergency approval of the BNT162b2 vaccine manufactured by Pfizer-BioNTech on December 11, 2020, vaccine administration began in Puerto Rico on December 15, 2020. The logistics of the vaccination process proposed by the Puerto Rico Department of Health established that the vaccine administration would be completed in phases, as recommended by the World Health Organization (WHO) and the Centers for Disease Control and Prevention (CDC).^10^ The phases (Supplementary Table S1) consisted on vaccinating populations that were at greater risk of infection and severe disease. For example, front line health care workers and people 65 years and older were vaccinated in the first phases.

By October 1, 2021 more than 2 million of the 3,285,874 individuals living in Puerto Rico had completed the COVID-19 vaccination series. Furthermore, since the start of the vaccination process to October 1, 2021, 111,052 cases have been detected with molecular or antigen tests in Puerto Rico with two surges, one starting in late March after restrictions were lifted and another in late June with the arrival of the Delta variant. We leveraged data collected by the Puerto Rico Department of Health to study the effect the vaccination process in preventing SARS-CoV-2 outcomes by comparing unvaccinated individuals 12 years or older to those who had completed the vaccination series (two weeks after final dose) for mRNA-1273, BNT162b2, or Ad26.COV2.S COVID-19 vaccines. In particular, we provide estimates of the effectiveness of the three COVID-19 vaccines protecting against infections, hospitalizations, and deaths to quantify the public-health impact of the vaccine in Puerto Rico.

## Methods

### Data sources

Two Puerto Rico Department of Health databases were integrated: the BioPortal, which stores test results, most hospitalizations, and deaths, and the Puerto Rico Electronic Immunization System (PREIS), which stores vaccination related data. For the analysis in this study we used daily counts of laboratory-confirmed SARS-CoV-2 infections, hospitalizations, and deaths from December 15, 2020 to October 1, 2021. We defined the first day to which an individual tested positive to a molecular or antigen test as the day of the infection. This same date was used for the analyses involving hospitalizations and deaths.

These daily counts were stratified by gender, age group (12-17, 18-24, 25-34, 35-44, 45-54, 55-64, 65-74, 75-84, or 85+) and vaccination status (unvaccinated, mRNA-1273, BNT162b2, or Ad26.COV2.S). Fully vaccinated was defined as 14 days after the final dose in the vaccine series. Cases in which the infection occurred after the first dose but before being fully vaccinated were removed from the analysis. Furthermore, the analysis was applied to two periods, before and after June 15, 2021, the approximate date in which the Delta variant became dominant in Puerto Rico.

### Statistical analyses

We applied models that accounted for age, gender, and, since many more tests were performed on weekdays than weekends, day of the week. We used the estimates obtained from fitting these models to quantify effectiveness and relative risks. The models used for the different analyses are described below.

### Time-varying effectiveness against infection by age, manufacturer

We defined 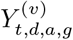 as the number of laboratory-confirmed SARS-CoV-2 infections observed on day *t* for individuals of gender *g* (male or female) and age group *a* (12-17, 18-24, 25-34, 35-44, 45-54, 55-64, 65-74, 75-84, or 85+), that were fully vaccinated with vaccine *v* (mRNA-1273, BNT162b2, or Ad26.COV2.S), *d* days previous to the positive test. We assumed these counts followed an over-dispersed Poisson distribution with expected rate defined by

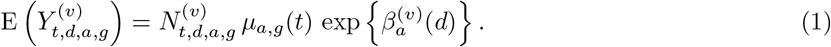

Here 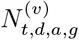 is the population size on day *t* of individuals of gender *g*, age group *a* that were fully vaccinated with vaccine *v, d* days before day *t, µ*_*a,g*_(*t*) is the incidence rate on date *t* among the non-vaccinated individuals of gender *g* and age *a*, and 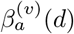 is the age-specific log relative risk of vaccine *v, d* days after the first dose was administered. Note the the transformation *f* (*x*) = 1 − exp(*x*) can be used to convert from relative risk to effectiveness. Since incidence changes across demographic groups and across days of the week (more tests performed on Mondays and less on Sundays, for example) we modeled the *µ*_*a,g*_(*t*) as

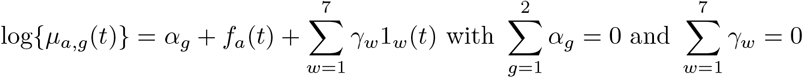

with *α*_*g*_ a gender effect, *f*_*a*_(*t*) a cubic-spline representing an age-specific trend that accounts for the change in incidence across time, and 1_*w*_(*t*) = 1 indicator functions for each day of the week *w* ∈ {Sunday, Monday, … Friday}. To estimate *µ*_*a,g*_(*t*) we assumed laboratory-confirmed SARS-CoV-2 infections counts for non-vaccinated individuals, denoted with 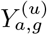, follow a Poisson distribution with rate *µ*_*a,g*_(*t*) and obtained the MLE for all parameters using Iteratively Reweighted Least Square algorithm, as implemented by the *glm* function in R. We then used these to define the MLE for *µ*_*a,g*_(*t*) and denoted it with 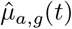. With this estimate in place we then estimated the parameter of interest 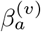 by treating *µ* _*a,g*_(*t*) in (1) as a known value, plugged in 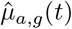, and noting that 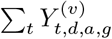 follows a Poisson distribution with expected value

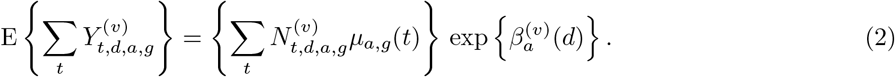

Because the first term on the right is treated as known, if we assume 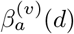 is a cubic-spline, then the model defined by (2) is reduced to a simple generalized linear model and we find the MLE 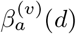 again using Iteratively Reweighted Least Square.

### Relative risk of hospitalization and death

Note that we can use this model for the hospitalization and deaths outcomes by simply redefining the counts 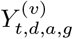 based on counts of these outcomes. Since we observed less hospitalization and death data than for infections, to increase power we also ran an analysis that assumed 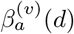 was constant across *d* and the same across age groups. Specifically we assumed that the age groups 18-44, 45-74, 75-84, and 85+ shared the same 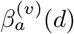. We then fit model (1) to obtain estimates for the relative risks.

### Hospitalization and death relative risk conditioned on infection

Note: in this section, to avoid introducing more mathematical symbols, we repurpose the *Y, N, α, β* and *γ* notation and, because we fit a model separately to each age group, we do not use the *a* index.

To estimate the further protection provided by the vaccine in reducing hospitalization and deaths among infected individuals we examined the proportion of cases that had hospitalizations or deaths. To do this we defined *N*_*i,t*_ as the number of laboratory-confirmed SARS-CoV-2 infections on day *t* for group *i*. Groups were defined by gender (male or female) and vaccination status (unvaccinated, mRNA-1273, or BNT162b2). Note that we did not include the Ad26.COV2.S vaccine in this analysis due to small sizes obtained after stratifying by age group. We defined *Y*_*i,t*_ as the number of these that had the hospitalization or death outcome. We then assumed that *Y*_*i,t*_ followed a binomial distribution with *N*_*i,t*_ trials and success probability *p*_*i,t*_ defined by:

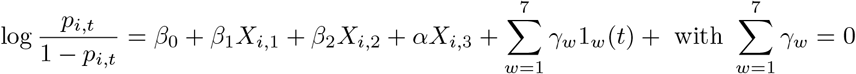

with *β*_0_ the baseline log odds for the unvaccinated, *X*_*i*,1_ = 1 if group *i* was vaccinated with Ad26.COV2.S and 0 otherwise, *X*_*i*,2_ = 1 if group *i* were vaccinated with mRNA-1273 and 0 otherwise, *X*_*i*,3_ = 1 if group *i* are male and 0 otherwise, and 1_*w*_(*t*) indicator functions as in (1). We fit this model using the Iteratively Reweighted Least Square algorithm. Note that the parameters of interest were *β*_1_ and *β*_2_.

## Results

### Vaccination data

As of October 1, 2021, 2,217,547 of the 3,285,874 individuals living in Puerto Rico had been fully vaccinated: 1,243,969 with BNT162b2, 844,065 with mRNA-1273, and 129,513 with Ad26.COV2.S. The daily COVID-19 vaccination rates were high during the first four months of the vaccination process, successfully vaccinating (at least one dose) 50% of the population eligible for inoculation at the time (1,442,459 of 2,848,293).^11^ However, as observed in previous vaccination campaigns, vaccine rates decreased after reaching 50%. The plateau was particularly noticeable among the older age groups. After several vaccine mandates were implemented in August and September, the rate commenced increasing (Figure 1). With the exception of pediatric population 12 years and older, whom were only vaccinated with BNT162b2, the age distribution of the vaccine was similar for all vaccine manufacturers (Supplementary Figure S1).

**Figure 1:**
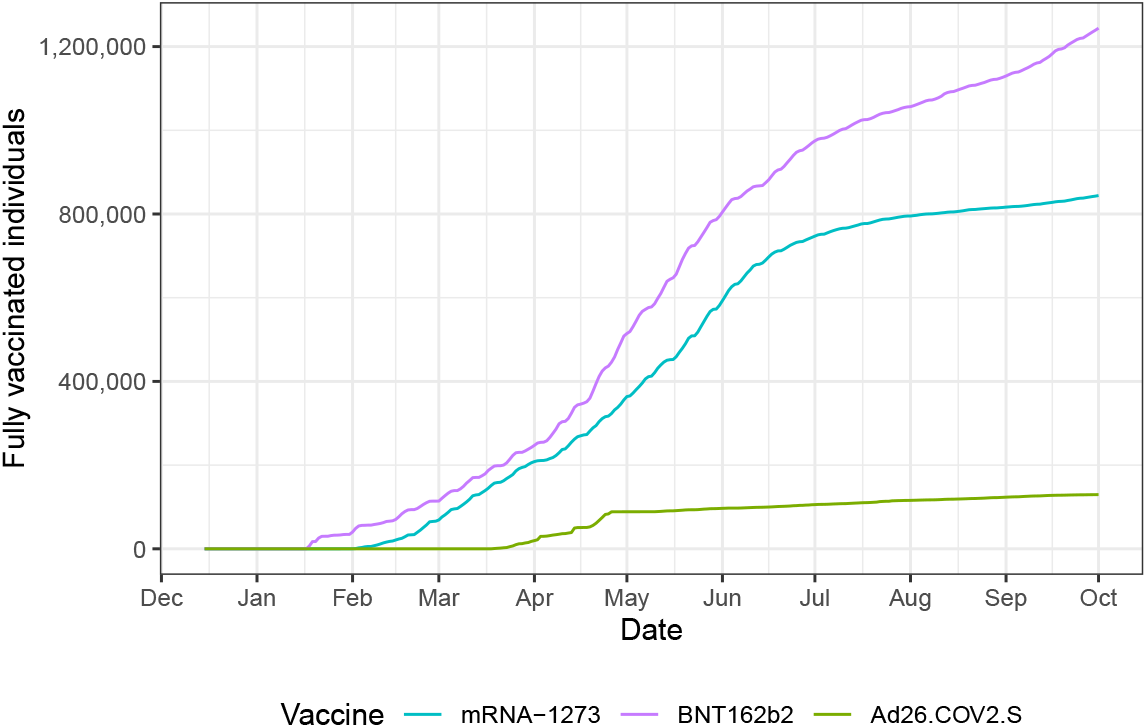
Total number of fully vaccinated individuals per day by vaccine manufacturer (represented with different colors).

### Observed cases, hospitalizations and deaths

During the period considered in this analysis (December 15, 2020 to October 1, 2021), 87,399 laboratory-confirmed SARS-CoV-2 infections were observed for individuals 12 years or older. These were mostly distributed across two surges (Figure 2). The first of these started shortly after several restrictions were lifted on March 15, 2021. Restrictions were imposed again on April 9, 2021 and the cases decreased to a level not seen since the summer of 2020. As a result, restrictions were lifted once again on May 24, 2021 and shortly after the arrival of the Delta, a second surge began by the end of June 2021.^12^ The SARS-CoV-2 infections observed during the period of December 15, 2020 to October 01, 2021 resulted in at least 4,942 and 1,333 COVID-19 related hospitalizations and deaths, respectively (Figure 2 and Supplementary Table S2).

**Figure 2:**
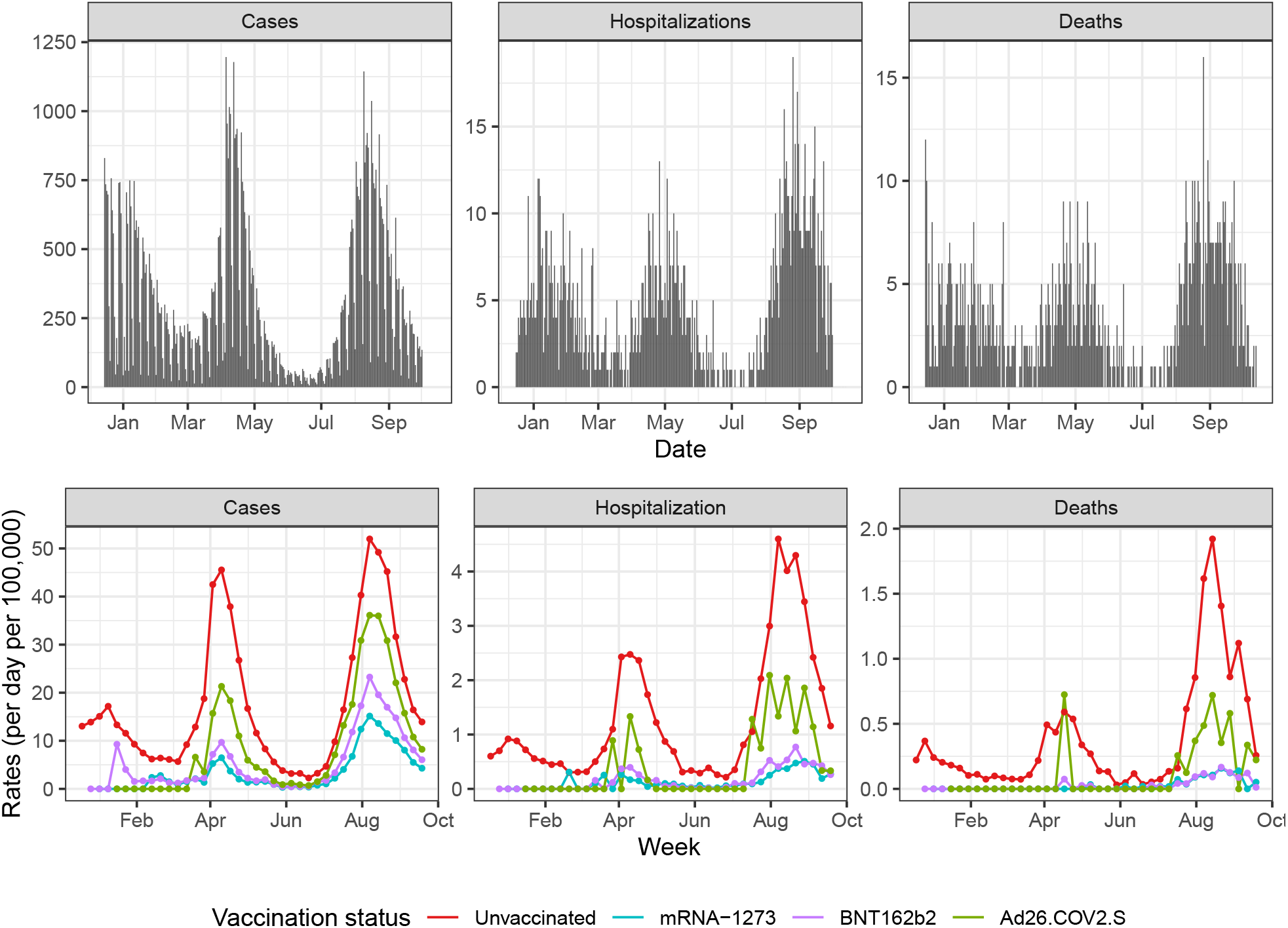
Above: Cases, hospitalizations and deaths per day for individuals 12 years or older during the period of study. Below: Weekly rates (per day per 100,000) for detected cases, hospitalizations, and deaths by vaccination status. Note that not all hospitalizations are reported to the BioPortal database. The rates are based on total numbers and are not adjusted for age. Colors are used to denote the different vaccination status.

The infections, hospitalization, and death rates were noticeably lower among the vaccinated (Supplementary Table S2). The protection afforded by the vaccine was observed by simply plotting the infection rates by vaccination group (Figure 2). The relative risk for the vaccinated seemed to increase in the period in which the Delta variant dominated. However, these rates are not age adjusted nor are potential effects of waning effectiveness considered. Therefore, to better assess COVID-19 vaccine effectiveness, we fit statistical models that account for time since the first dose, age, gender, and other potential confounding variables.

### Time-varying vaccine effectiveness against infections

We fit a statistical model (described in detail in the Methods Section) to estimate time-varying effectiveness while accounting for the difference in demographics between vaccinated and unvaccinated groups. Note that vaccine administration for individuals between 12 and 17 years started on May 12, 2021 and only with the BNT162b2 vaccine.

To evaluate whether the Delta variant affected vaccine effectiveness, we considered two periods: before and after June 15, 2021. The mRNA-1273 and BNT162b2 vaccines remained steady at over 75% effective after the Delta variant gained dominance and a decrease in vaccine effectiveness was not detected due to the Delta variant (Supplementary Figure S2).

Furthermore, to evaluate if there were differences in effectiveness across age groups for the mRNA-1273 and BNT162b2 vaccines we ran a similar analysis, but comparing age groups. We did not observe substantial differences in effectiveness against infections by age, except for those 85 and older for whom the waning appears worse (Supplementary Figure S3). Note that this group was smaller and resulted in less precise estimates.

Considering that no clear differences were observed across age groups and time-periods, we estimated one time-varying effectiveness across all age-groups and for the entire vaccination process period. Note that the period of evaluation was different for the different age groups given the previously described phases (Supplementary Table S1). Furthermore, for the Ad26.COV2.S vaccine, the period after the first dose administration date was shorter because this vaccine received its EUA on March 3, 2021.

The mRNA-1273, BNT162b2, and Ad26.COV2.S vaccines had peak effectiveness of 90% (88% - 91%), 87% (85% - 89%), and 58% (51% - 65%), respectively (Figure 3). Effectiveness then commenced waning to 71% (68% - 74%), 56% (53% - 59%), and 27% (17% - 37%), respectively, at the end of the study period (Supplementary Table S3).

**Figure 3:**
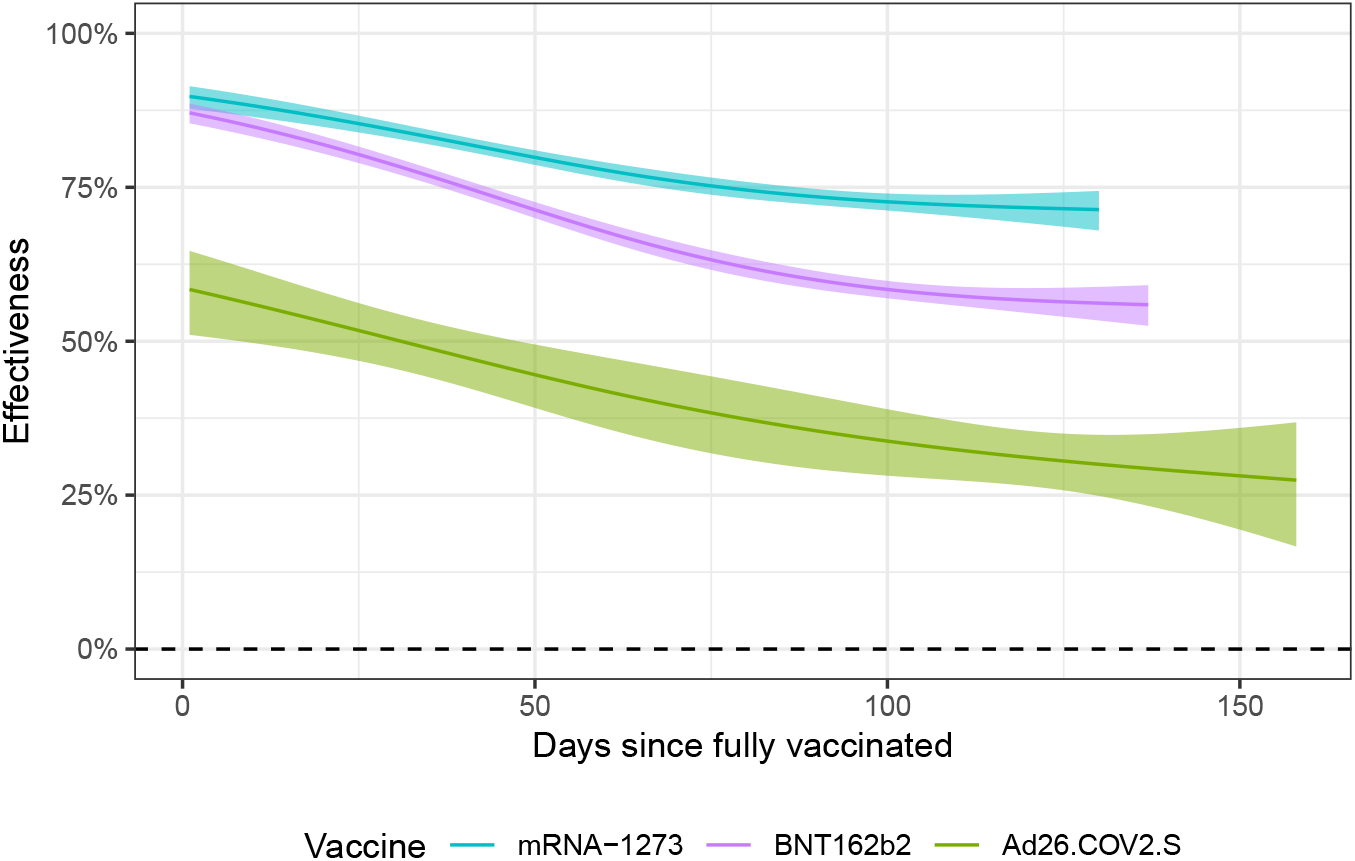
Vaccine effectiveness through time. The estimated effectiveness is plotted against days since the individuals were fully vaccinated. The ribbons represent point-wise 99% confidence intervals. The different vaccine manufacturers are denoted in different colors.

We fit the same model to estimate relative risk of hospitalization comparing vaccinated and unvaccinated individuals across time since fully vaccinated. Although we did not have enough data to obtain as precise estimates as for infections, similar patterns were observed (Supplementary Figure S4).

### Vaccines lower the risk of hospitalization and death

We fit a statistical model (described in detail in the Methods Section) to determine if the vaccines provided further protection against hospitalization and death among infected individuals. Specifically, we estimated the probability of hospitalization and death conditioned on individuals testing positive for SARS-CoV-2 for several age groups (Figure 4 and Supplementary Table S4). Due to the small sample sizes available for the Ad26.COV2.S vaccine, after stratification by age, we did not include it in this analysis. The probability for hospitalization after infection for those that were vaccinated was at least twice lower for all age groups except the 85 and older for which the probability was over 50% lower. Similarly, the probability of death after infection was over three times lower for all age groups, except the 85 and older for which it was about twice as low.

**Figure 4:**
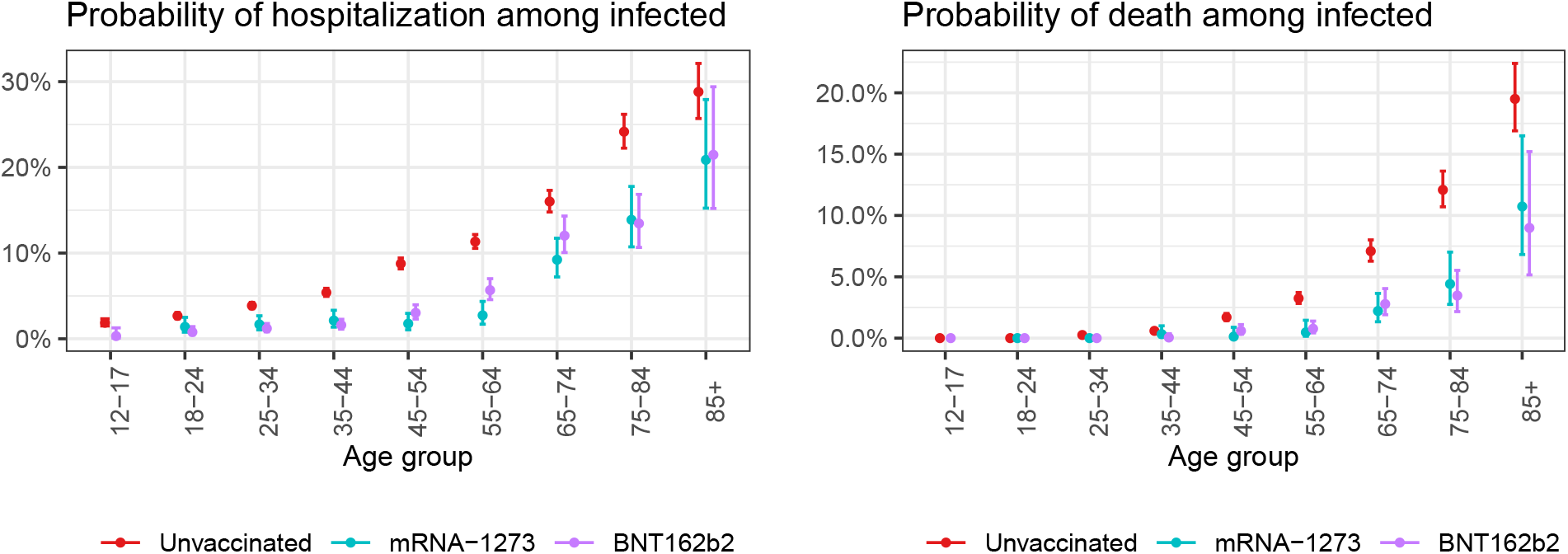
Estimated probability of hospitalizations and deaths from COVID-19 among individuals infected with SARS-CoV-2 with 95% confidence intervals. Two vaccinated groups are compared to unvaccinated with the different vaccination manufacturers denoted with color. Note that not all hospitalizations are reported to the BioPortal database.

We also fit a statistical model to estimate the relative risk of COVID-19 hospitalization and death comparing vaccinated and non-vaccinated individuals (not conditioned on being infected). Due to sparsity in the data we grouped age groups as described in the Methods Section. Individuals vaccinated with mRNA-1273 or BNT162b2 between 45 and 74 years were found to be 18 (15, 22) and 8.0 (7, 9.2) times less likely to be hospitalized due to COVID-19 in comparison to unvaccinated individuals, respectively. Similarly, Ad26.COV2.S vaccinated individuals between 45 and 74 years had 3.9 (3, 5.0) times less risk for hospitalizations in comparison to the unvaccinated. For those 75 and older vaccinated with mRNA-1273 or BNT162b2, their risk for hospitalization by COVID-19 is reduced by 2.8 (2, 4.0) and 2.9 (2, 4.2) times, respectively, in comparison to unvaccinated individuals. We did not find statistically significant evidence that the Ad26.COV2.S vaccine provided added protection against hospitalization for people 85 and older (Table 1).

**Table 1:**
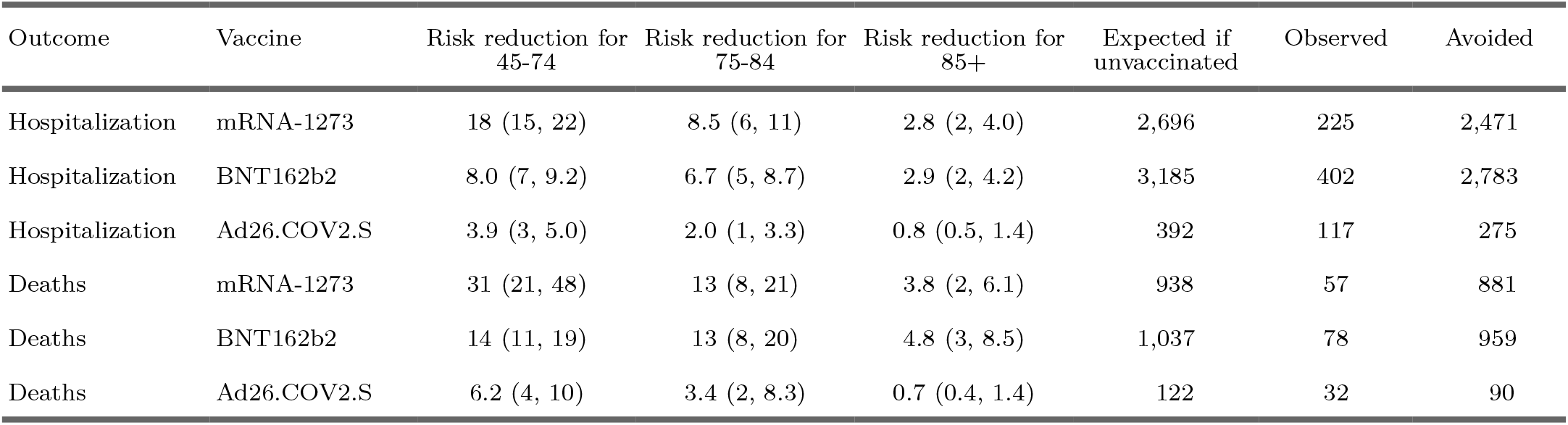
Reduction in risk provided by the different vaccines for individuals across age groups. We estimated the relative risk of hospitalization and death for these two age groups and each vaccine manufacturer, with a model that adjusted for population size, age, gender, and time varying incidence rates. Columns 3, 4, and 5 show estimated reduction in risk and 95% confidence intervals, comparing unvaccinated to vaccinated rates. Columns 6, 7, and 8 show estimated expected outcomes based on estimated unvaccinated rates and the difference between these.

All three vaccines provided high protection against death by COVID-19 for those between 45 and 84 years. For people 85 years vaccinated with mRNA-1273 or BNT162b2, the risk reduction was 3.8 (2, 6.1) and 4.8 (3, 8.5), respectively, showing these vaccines provide great protection against death for this age group. The Ad26.COV2.S vaccine did not provide added protection against death by COVID-19 for people 85 and older (Table 1).

## Discussion

Our analyses clearly demonstrates that all vaccines were effective at reducing risks of infection, hospitalization and death across all age groups, with the protection provided by the and mRNA-1273 and BNT162b2 vaccine particularly strong. At the peak of their effectiveness mRNA-1273 and BNT162b2 were 90% (88% - 91%) and 87% (85% - 89%) effective, respectively. Meanwhile Ad26.COV2.S was 58% (51% - 65%) at its peak. The effectiveness of all vaccines decreased over tine, although mRNA-1273’s effectiveness remained high at 71% even after four months. BNT162b2 was also relatively high, at around 56%, however, Ad26.COV2.S effectiveness dropped to 27%. All vaccines had a lower effectiveness for those over 85 years, with the decrease in effectiveness particularly low for the Ad26.COV2.S vaccine. No evidence was found that effectiveness was different after the Delta variant became dominant.

It is important to note that this study is observational based on real world data and not a randomized clinical trial. Although we controlled for confounding factors that were coded and available for analyses, it is possible that there were unknown and unobserved confounders that were not accounted for, such as chronic health conditions and immunocompromising conditions. However, the findings described here agree with general trends observed in the clinical trials.^2 3^.^4^

Following CDC guidelines, in Puerto Rico it is general practice to recommend diagnostic testing to those who are experiencing symptoms and/or are direct contacts of a case.^13^ This implies there is a risk for selection bias given the fact that vaccinated people are less likely to experience severe illness or symptoms, and thus less likely to get a diagnostic test.^14^

It is also important to note that to perform this analysis two Puerto Rico Department of Health databases were integrate: the BioPortal, which stores test results, hospitalizations, and deaths, and PREIS, which stores vaccination data. These two databases lacked a common unique identifier and records were matched using patient information. This implies that some parings may have been missed, although a human spot check found the paring to be 99% accurate. We also note that due to deficiencies in reporting within the Department of Health, not all hospitalizations and deaths were included in the BioPortal database and there is lag in reporting in some vaccination centers. However, no clear reason that could bias results to lower or higher vaccination effectiveness estimates was detected. Finally, it is also important to note that all our analyses depend on age specific population estimates provided by the United States Census Bureau and these are likely to change with the finalization of the 2020 census.

## Data Availability

The code and data needed to reproduce the analysis is made publicly available here: https://github.com/rafalab/vax-eff-pr

## Acknowledgements

We thank the Puerto Rico Department of Health and Digheontech, Inc. for allowing us to collaborate in this effort. In particular, we thank doctors Melissa Marzán, and Daniel Colón-Ramos for their continued support and suggestions in the completion of this project. Furthermore, we thank Wilmarí de Jesús Álvarez and Dr. Fabiola Cruz López for providing insight into aspects concerning demography and molecular epidemiology, respectively.

## Supplementary Material

### Supplementary Figures

**Figure S1:**
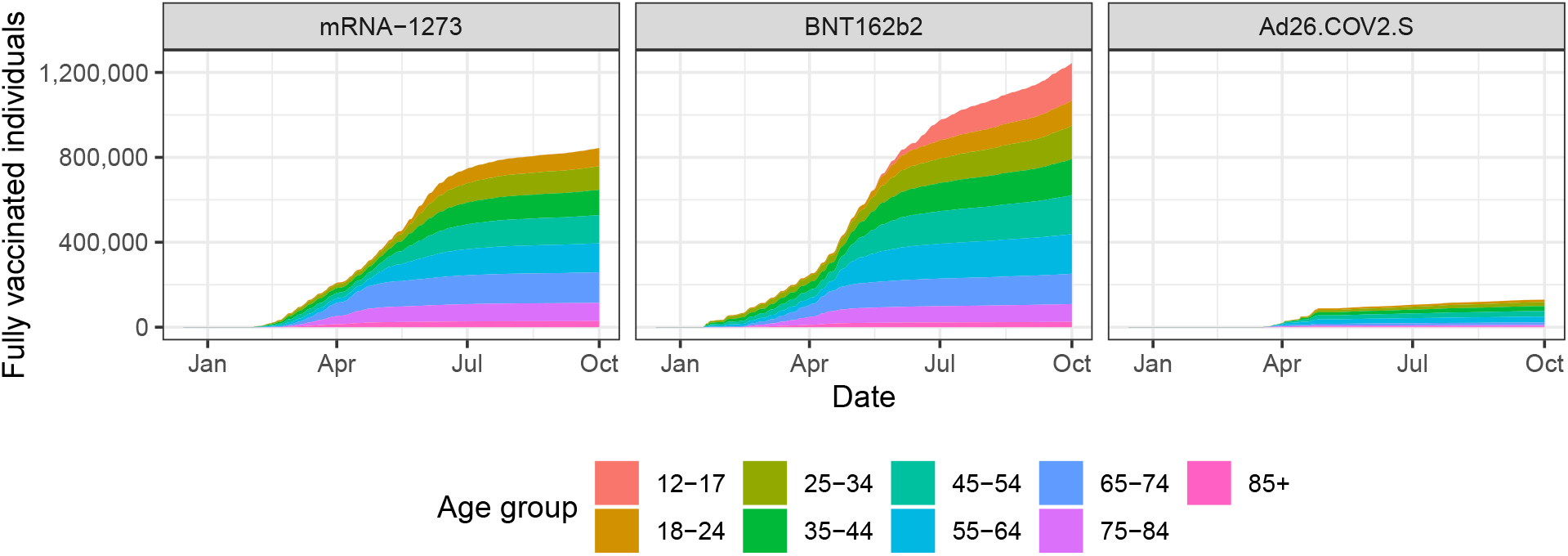
Total number of fully vaccinated individuals by age group and vaccine manufacturer. Each pane represents a vaccine manufacturer and colors represent the different age groups.

**Figure S2:**
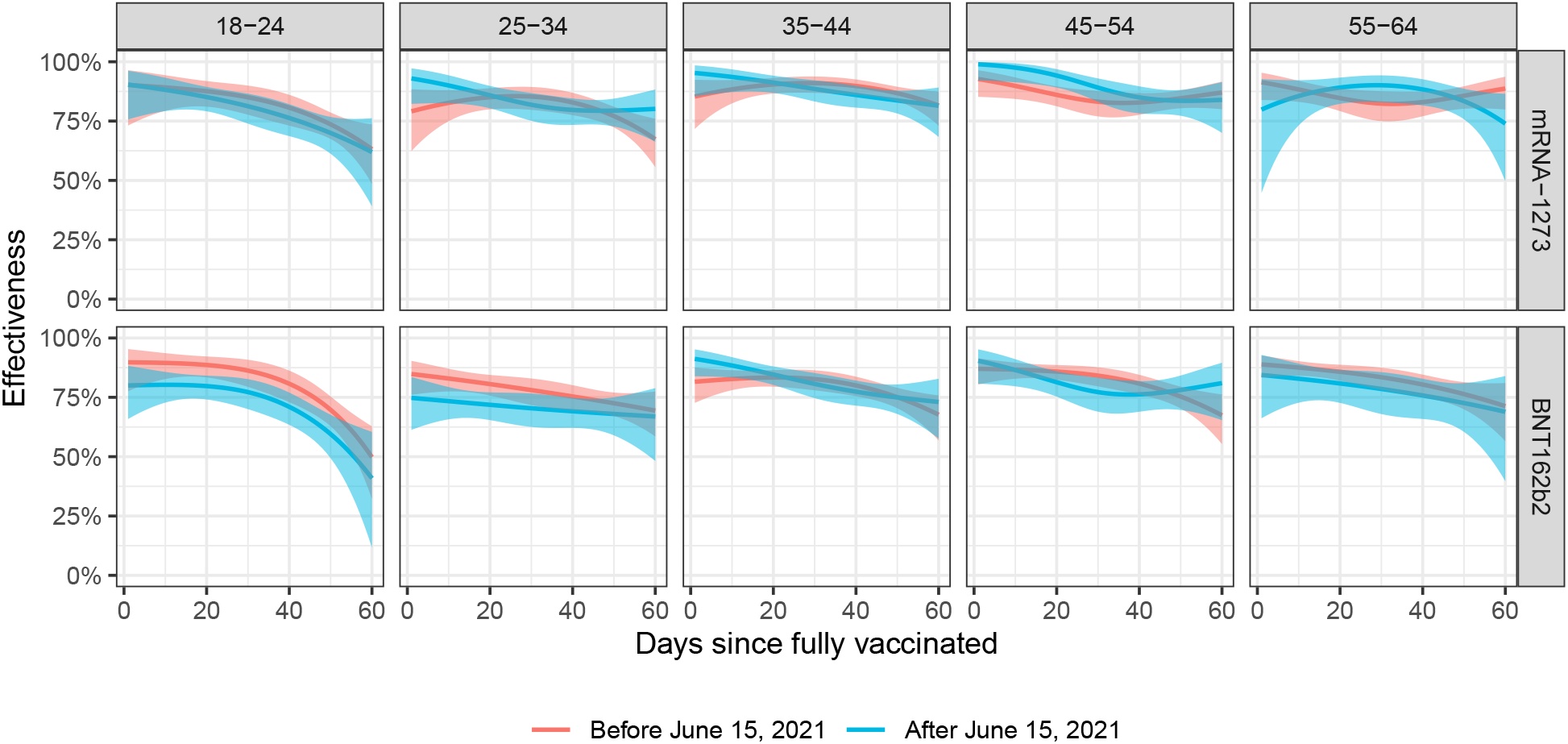
Time varying effectiveness estimates by age group and vaccine manufacturer before and after arrival of the Delta variant. The ribbons represent point-wise 99% confidence intervals.

**Figure S3:**
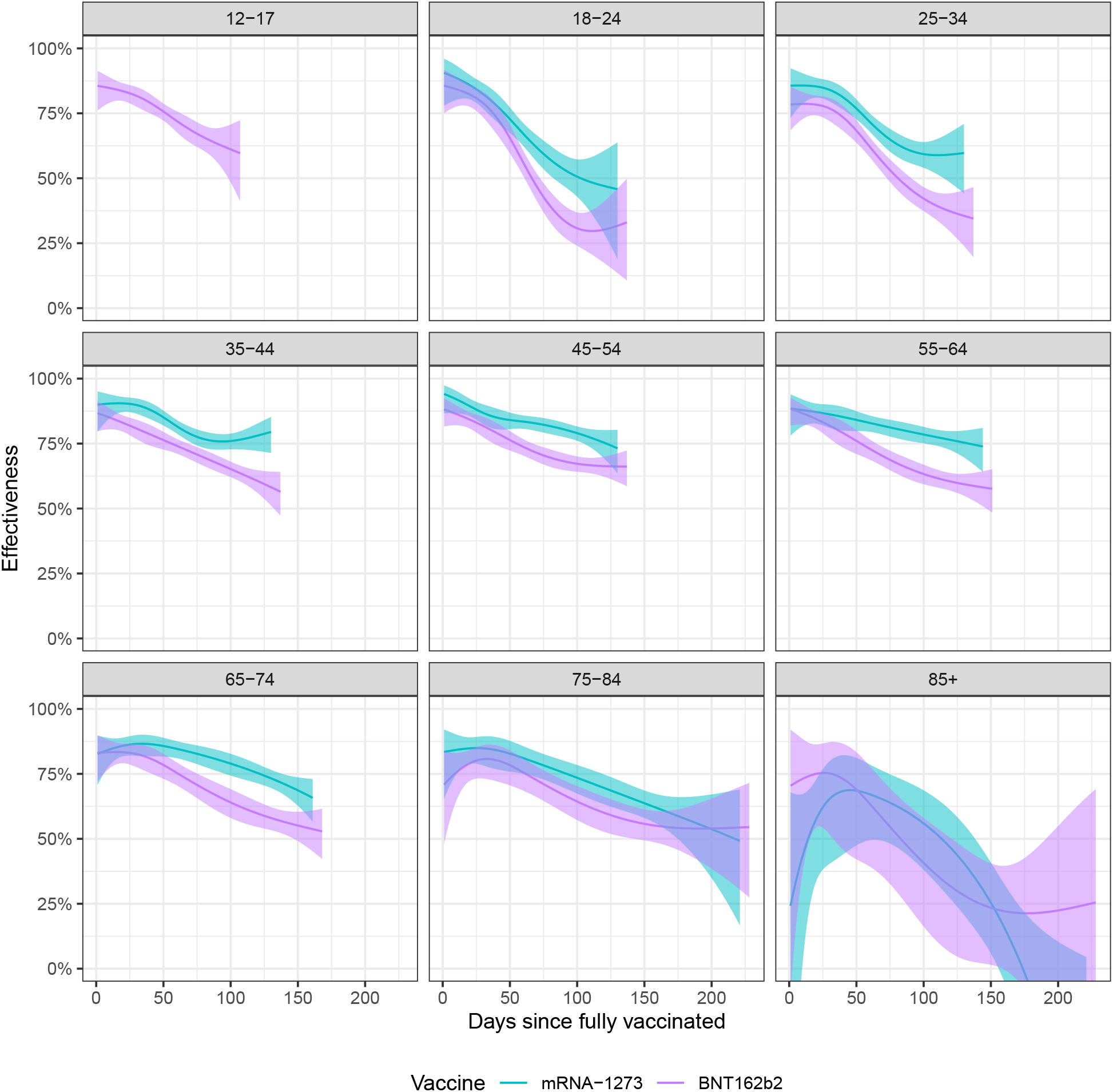
Waning effectiveness against infection by age group and vaccine manufacturer (colors). The ribbons represent point-wise 99% confidence intervals.

**Figure S4:**
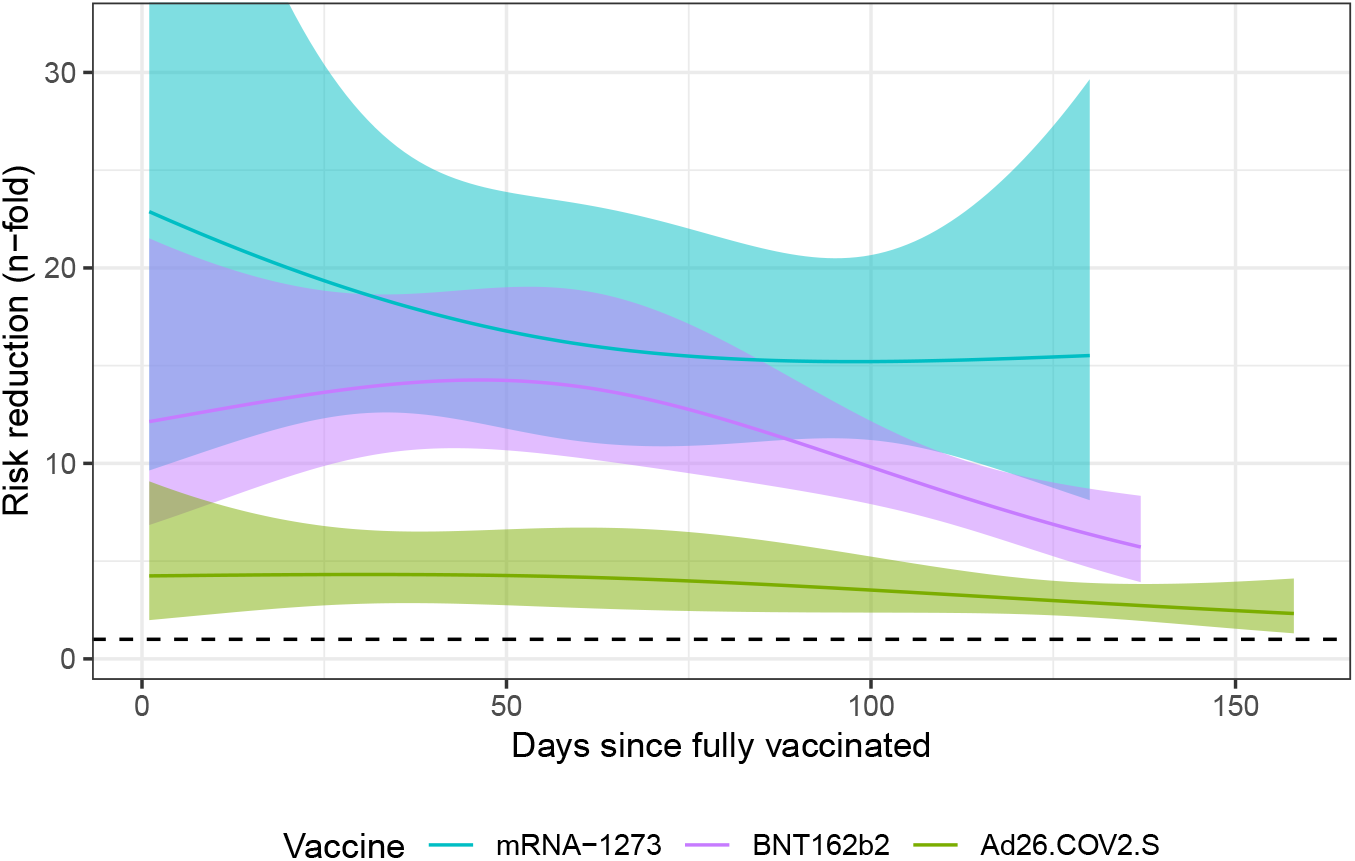
Vaccine reduction in risk of hospitalization through time. The estimated effectiveness is plotted against days since the first dose was administered. The different vaccine manufacturers are denoted in different colors. The ribbons represent point-wise 99% confidence intervals.

### Supplementary Tables

**Table S1:** Chronology of the COVID-19 vaccine rollout in Puerto Rico.

| Date | What happened? |
| --- | --- |
| December 15, 2020 | The COVID-19 vaccine rollout begins with phase 1A which included healthcare workers, communities in long term care facilities and intellectual disabilities care facilities. |
| December 18, 2020 | mRNA-1273 (Moderna) receive Emergency Use Authorization (EUA). |
| January 11, 2021 | Phase 1B commences with the vaccination of adults 65 and older. |
| February 2, 2021 | Puerto Rico's Secretary of Health signs administrative order establishing that for the next 28 days, vaccination will be exclusive for those 65 and older. |
| March 3, 2021 | Ad26.COV2.S (Johnson & Johnson) receives EUA |
| March 11, 2021 | On March 3rd, 2021 Secretary of Health signs administrative order establishing that starting on March 11th and for the following 30 days, first doses are to be administered exclusively to adults 60 and older with certain chronic conditions. |
| March 12, 2021 | On March 10th, 2021, Secretary signs administrative order establishing that starting on March 11th and for the following 30 days, first doses are to be administered exclusively to adults 60 and older and 50 to 59 year olds with chronic conditions. |
| March 17, 2021 | Secretary of Health signs executive order authorizing the vaccination of personnel in food industry, drug companies, medical equipment, the public transport sector, air transport and maritime cargo. |
| March 29, 2021 | Phase 1C begins with the vaccination of people 50 and older and 35 and older with chronic conditions. |
| April 12, 2021 | Phase 2 begins with vaccination available to everyone 16 and older. |
| May 12, 2021 | CDC recommends vaccination for people 12 years and older. |

**Table S2:**
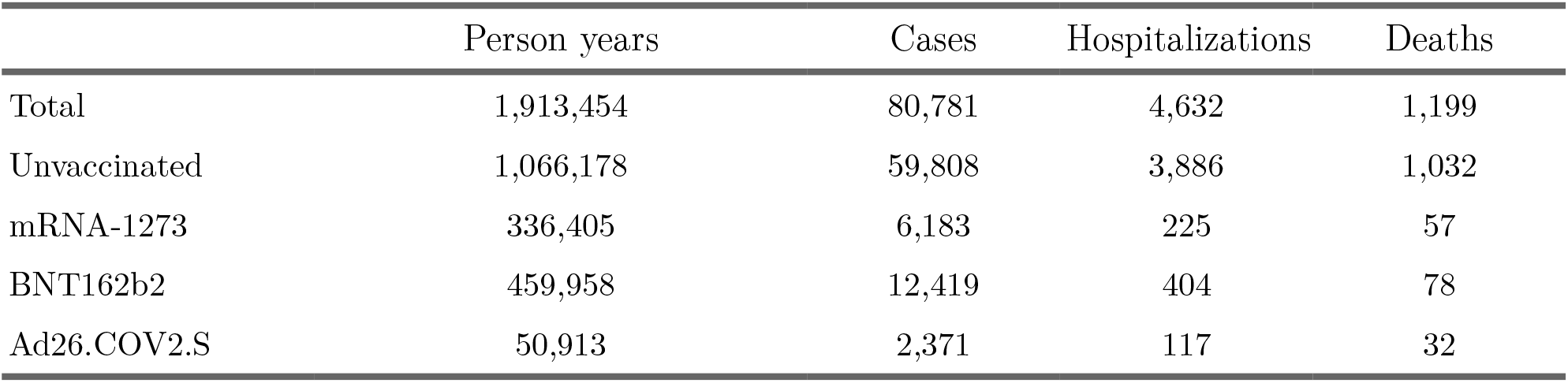
Person years, infections, hospitalizations, and deaths from December 29, 2020, the first day that individuals were fully vaccinated, to October 1, 2021by vaccination group.

|  | Person years | Cases | Hospitalizations | Deaths |
| --- | --- | --- | --- | --- |
| Total | 1,913,454 | 80,781 | 4,632 | 1,199 |
| Unvaccinated | 1,066,178 | 59,808 | 3,886 | 1,032 |
| mRNA-1273 | 336,405 | 6,183 | 225 | 57 |
| BNT162b2 | 459,958 | 12,419 | 404 | 78 |
| Ad26.COV2.S | 50,913 | 2,371 | 117 | 32 |

**Table S3:** Waning effectiveness against infection with 99% point-wise confidence intervals. The periods examined due to the fact that different vaccines were approved at different times and have different times since first dose to being fully vaccinated.

| Vaccine | Peak effectiveness (CI) | Length of period examined | Effectiveness (CI) at end of period |
| --- | --- | --- | --- |
| mRNA-1273 | 90% (88% - 91%) | 130 days | 71% (68% - 74%) |
| BNT162b2 | 87% (85% - 89%) | 137 days | 56% (53% - 59%) |
| Ad26.COV2.S | 58% (51% - 65%) | 158 days | 27% (17% - 37%) |

**Table S4:** Estimated probability of hospitalizations and deaths from COVID-19 among individuals infected with SARS-CoV-2. Note that the mRNA-1273 vaccine is not approved for individuals younger than 18. Thus no probabilities are reported for the 12-17 age group for this vaccine.

| Outcome | Age group | Unvaccinated | BNT162b2 | mRNA-1273 |
| --- | --- | --- | --- | --- |
| Hospitalization | 12-17 | 0.02 (0.02,0.02) | 0.00 (0.00,0.01) | - |
| Hospitalization | 18-24 | 0.03 (0.02,0.03) | 0.01 (0.00,0.01) | 0.01 (0.01,0.03) |
| Hospitalization | 25-34 | 0.04 (0.04,0.04) | 0.01 (0.01,0.02) | 0.02 (0.01,0.03) |
| Hospitalization | 35-44 | 0.05 (0.05,0.06) | 0.02 (0.01,0.02) | 0.02 (0.01,0.03) |
| Hospitalization | 45-54 | 0.09 (0.08,0.09) | 0.03 (0.02,0.04) | 0.02 (0.01,0.03) |
| Hospitalization | 55-64 | 0.11 (0.11,0.12) | 0.06 (0.05,0.07) | 0.03 (0.02,0.04) |
| Hospitalization | 65-74 | 0.16 (0.15,0.17) | 0.12 (0.10,0.14) | 0.09 (0.07,0.12) |
| Hospitalization | 75-84 | 0.24 (0.22,0.26) | 0.13 (0.11,0.17) | 0.14 (0.11,0.18) |
| Hospitalization | 85+ | 0.29 (0.26,0.32) | 0.21 (0.15,0.29) | 0.21 (0.15,0.28) |
| Deaths | 12-17 | 0.00 (0.00,0.00) | 0.00 (0.00,0.00) | - |
| Deaths | 18-24 | 0.00 (0.00,0.00) | 0.00 (0.00,0.00) | 0.00 (0.00,0.00) |
| Deaths | 25-34 | 0.00 (0.00,0.00) | 0.00 (0.00,0.00) | 0.00 (0.00,0.00) |
| Deaths | 35-44 | 0.01 (0.00,0.01) | 0.00 (0.00,0.00) | 0.00 (0.00,0.01) |
| Deaths | 45-54 | 0.02 (0.01,0.02) | 0.01 (0.00,0.01) | 0.00 (0.00,0.01) |
| Deaths | 55-64 | 0.03 (0.03,0.04) | 0.01 (0.00,0.01) | 0.00 (0.00,0.01) |
| Deaths | 65-74 | 0.07 (0.06,0.08) | 0.03 (0.02,0.04) | 0.02 (0.01,0.04) |
| Deaths | 75-84 | 0.12 (0.11,0.14) | 0.03 (0.02,0.06) | 0.04 (0.03,0.07) |
| Deaths | 85+ | 0.20 (0.17,0.22) | 0.09 (0.05,0.15) | 0.11 (0.07,0.16) |

## References

1. Pardi N, Hogan MJ, Porter FW, Weissman D. mRNA vaccines—a new era in vaccinology. Nature reviews Drug discovery 2018;17(4):261–79.

2. Polack FP, Thomas SJ, Kitchin N, et al. Safety and Efficacy of the BNT162b2 mRNA Covid-19 Vaccine. New England Journal of Medicine [Internet] 2020 [cited 2021 Sep 13];383(27):2603–15. Available from: https://doi.org/10.1056/NEJMoa2034577

3. Baden LR, El Sahly HM, Essink B, et al. Efficacy and Safety of the mRNA-1273 SARS-CoV-2 Vaccine. New England Journal of Medicine [Internet] 2021 [cited 2021 Sep 13];384(5):403–16. Available from: https://doi.org/10.1056/NEJMoa2035389

4. Sadoff J, Gray G, Vandebosch A, et al. Safety and Efficacy of Single-Dose Ad26.COV2.S Vaccine against Covid-19. New England Journal of Medicine [Internet] 2021 [cited 2021 Sep 13];384(23):2187–201. Available from: https://doi.org/10.1056/NEJMoa2101544

5. Aran D. Estimating real-world COVID-19 vaccine effectiveness in Israel. 2021 [cited 2021 Sep 13];2021.02.05.21251139. Available from: https://www.medrxiv.org/content/10.1101/2021.02.05.21251139v1

6. Dagan N, Barda N, Kepten E, et al. BNT162b2 mRNA Covid-19 Vaccine in a Nationwide Mass Vaccination Setting. New England Journal of Medicine [Internet] 2021 [cited 2021 Sep 13];384(15):1412–23. Available from: https://doi.org/10.1056/NEJMoa2101765

7. Thompson MG, Stenehjem E, Grannis S, et al. Effectiveness of COVID-19 vaccines in ambulatory and inpatient care settings. New England Journal of Medicine 2021;

8. Lopez Bernal J, Andrews N, Gower C, et al. Effectiveness of Covid-19 Vaccines against the B.1.617.2 (Delta) Variant. New England Journal of Medicine [Internet] 2021 [cited 2021 Sep 13];385(7):585–94. Available from: https://doi.org/10.1056/NEJMoa2108891

9. Nasreen S, Chung H, He S, et al. Effectiveness of COVID-19 vaccines against variants of concern in Ontario, Canada. 2021 [cited 2021 Sep 13];2021.06.28.21259420. Available from: https://www.medrxiv.org/content/10.1101/2021.06.28.21259420v2

10. COVID-19 vaccination program operational guidance [Internet]. Available from: https://www.cdc.gov/vaccines/covid-19/covid19-vaccination-guidance.html

11. Department of Health PR. COVID-19 EN CIFRAS EN PUERTO RICO [Internet]. [cited 2021 Sep 13];Available from: https://covid19datos.salud.gov.pr/www.salud.gov.pr

12. Universidad Central del Caribe YU. Informes de Proyecto Vigilancia Genómica [Internet]. [cited 2021 Sep 13];Available from: https://drive.google.com/drive/folders/1HcMGwEieXWp728PZOeUxQeZjWk_sf1a8

13. Overview of Testing for SARS-CoV-2 (COVID-19) [Internet]. 2020 [cited 2021 Sep 13];Available from: https://www.cdc.gov/coronavirus/2019-ncov/hcp/testing-overview.html

14. Disease Control and Prevention C for. The Possibility of COVID-19 after Vaccination: Breakthrough Infections [Internet]. 2021 [cited 2021 Sep 13];Available from: https://www.cdc.gov/coronavirus/2019-ncov/vaccines/effectiveness/why-measure-effectiveness/breakthrough-cases.html

